# Disentangling Fatigue from Depression among Survivors of Severe COVID-19

**DOI:** 10.64898/2026.04.24.26351694

**Authors:** Juan R. Cabrera, Peter Pham, W. John Boscardin, Anil N. Makam

## Abstract

**Purpose:** Survivors of severe COVID-19 commonly experience post-intensive care syndrome (PICS), which includes depression and fatigue. Fatigue is far more common and may inflate depression severity given overlapping symptoms. We sought to disentangle fatigue from depression in PICS.

**Methods:** We conducted a cross-sectional analysis of the RAFT COVID study, a national multicenter longitudinal cohort of severe prolonged COVID-19 survivors. We included participants who completed validated surveys at 1-year from hospitalization for depression (PHQ-9) and fatigue (FACIT-Fatigue). We described correlation of FACIT-fatigue with the PHQ9, and separately with PHQ-2 and PHQ-7, which both omit the two items we hypothesized are influenced by fatigue—tiredness and sleeping. Using a MIMIC model, we performed differential item functioning to evaluate the impact of fatigue on depression directly through these two questions and indirectly with the latent depression construct. We then compared PHQ-7 to PHQ-9 scores by fatigue status.

**Results:** Among 82 participants, 61.0% reported fatigue (reverse-scored FACIT-Fatigue ≥9), and 15.9% moderately severe depression (PHQ-9 ≥10). FACIT-fatigue was strongly correlated with PHQ-9 (r=.87, p<.001), but less so for PHQ-2 (r=.76, p<.001) and PHQ-7 (*r=*.82, p<.001). The MIMIC model identified significant direct effects on tiredness (λ=.89, p<.001) and sleep (λ=.52, p<.001). Among fatigued participants, the rescaled PHQ-7 was lower than the PHQ-9 (median of 4.5, IQR 1.50-9.75, vs 7, IQR 4-9.75).

**Conclusions:** Fatigue significantly inflated depression symptoms in severe COVID-19 survivors through tiredness and sleeping PHQ-9 items. PHQ-2 may better screen for true depressive symptoms in PICS, minimizing the risk of misdiagnosis and overtreatment.

**PLAIN ENGLISH SUMMARY:** Survivors of severe COVID-19 illness commonly experience post-intensive care syndrome (PICS), which includes depression and fatigue. Fatigue is far more common and may inflate depression severity given overlapping symptoms. We sought to disentangle fatigue from depression in PICS. We found that the presence of fatigue inflated depression severity through symptoms of tiredness and difficulty sleeping, which are two of the nine items of a commonly used depression screening tool, known as the Patient Health Questionnaire-9 (PHQ-9). Depression screening tools that omit these two items, such as the PHQ-2, may better screen for depressive symptoms in PICS, minimizing the risk of overestimating depression symptoms and potentially misdiagnosis.

## INTRODUCTION

The pandemic has led to a surge of survivors of severe respiratory failure from COVID-19 due to pneumonia and acute respiratory distress syndrome (ARDS).^1–3^ Based on what we know from decades of research on survivors of ARDS and critical illness,^4–11^ as well as from the evolving literature on severe COVID-19,^12–20^ survivors of severe COVID-19 are at high risk of developing persistent impairments known as post-intensive care syndrome (PICS), and potentially also Long COVID. One of the most prominent features of both PICS and Long COVID is depression, which occurs in about 15-20% after a severe COVID illness.^12,14,19^

One major challenge in identifying survivors of severe COVID-19 who may be eligible for depression treatment in clinical practice and in assessing the prevalence of depression for epidemiologic research is disentangling depression from fatigue. This is because fatigue is a well-recognized symptom of depression and is included in the Patient Health Questionnaire-9 (PHQ-9),^21^ a commonly used screening tool for depression, and is also highly prevalent after severe COVID-19 illness, occurring in up to 60% of survivors.^12,19,22^ Thus, clinicians and researchers may overestimate the burden and severity of depression in the setting of fatigue after severe COVID-19.

Current studies that examined PICS and long COVID have not disentangled depression from fatigue. However, previous studies of other illnesses where both of these symptoms commonly cooccur, including multiple sclerosis and chronic fatigue syndrome,^23–25^ have found that fatigue inflates the prevalence and severity of depression based on screening tools like the PHQ-9. Distinguishing depression from fatigue in severe COVID-19 survivors can better inform the burden of depression after severe COVID-19 and guide more appropriate diagnosis and treatment.

Therefore, in this study, we sought to distinguish depression from fatigue among adults who participated in the Recovery After Transfer to a Long-Term Acute Care Hospital (LTACH) for COVID-19 (RAFT COVID)—a national multicenter prospective study on 1-year outcomes after prolonged severe COVID-19 illness.^12^ We hypothesized that we would also find a significant overlap between fatigue and depression after severe COVID-19, supporting the need to use alternate depression screening scales and corroborate its presence and severity with more in-depth psychiatric assessments.

## METHODS

### Study Design and Study Sample

We conducted a cross-sectional analysis of the Recovery After Transfer to an LTACH for COVID-19 (RAFT COVID) study, which was a national nine center prospective longitudinal cohort of English-speaking patients with among the most severe and prolonged COVID-19 illness between March 2020 and February 2021 who survived at least 1 year from the LTACH admission date.^12^ Each participating LTACH maintained an active list of COVID-19 admissions, which was confirmed by clinical study site personnel by reviewing the medical record to both confirm evidence of COVID-19 illness upon admission and ensure that we did not include incidental SARS-CoV-2 or LTACH-acquired infection. Personnel at each LTACH attempted three phone calls after discharge to obtain permission to be contacted by a research team member. Among assenting participants, a research team member attempted up to three phone call attempts and two e-mails (if provided). We added an assessment of fatigue midway during the study (September 9, 2021) given evolving evidence that COVID-19 survivors had increased fatigue. For this study, we included all participants 18 years or older who completed the depression and fatigue validated questionnaires during a telephone-administered or web-based survey at the one-year follow-up visit from the date of the index acute care hospitalization preceding the LTACH stay. The University of California, San Francisco Institutional Review Board approved the RAFT COVID study on June 18, 2020 (#20-31060). Further details of the RAFT COVID study have been published.^12^

### Characteristics of the Cohort

We collected information on sociodemographic status, baseline health status, severity of illness, and one-year health status spanning functioning, cognition, psychiatric symptoms, and quality of life domains using: a) participant self-report from the survey; b) LTACH Continuity Assessment Record and Evaluation, Version 4.0 (from the Centers for Medicare and Medicaid Services, Baltimore, MD),^26^ which is a federally mandated assessment on admission and discharge; and c) medical chart review by trained clinical staff at each LTACH entered into a web-based case record form.

### Fatigue Assessment

We used the Functional Assessment of Chronic Illness Therapy (FACIT)-Fatigue scale, which is a validated instrument comprising 13 items that participants rate based on their experiences over the past one week.^27^ Responses are scored on a 5-point Likert scale, ranging from 0 (“not at all”) to 4 (“very much”). To facilitate interpretation of our model findings, we reverse-scored the scale such that higher scores indicated greater fatigue to parallel the directionality of the depression scale. We reverse scored FACIT-Fatigue, which ranges from 0-52, so that higher scores indicated greater fatigue to parallel the directionality of the depression scale. Thus, a score of ≥9 on the reverse-scored scale was defined as fatigued (which corresponded to ≤43 cutoff on the standard scale which was identified as the optimal cutpoint to differentiate fatigue in the disease state of anemia from cancer versus a general population).^28^

### Depression Assessment

We used the Patient Health Questionnaire-9 (PHQ-9) to screen for the presence and assess the severity of depressive symptoms, a validated and widely-used instrument consisting of 9 items, each corresponding to the diagnostic criteria for major depressive disorder per the DSM-V.^21^ Respondents are asked to indicate how often they have been bothered by each symptom over the past two weeks, with response options scored from 0 (“not at all”) to 3 (“nearly every day”). The total score ranges from 0 to 27, where higher scores signify greater severity of depression. A score of ≥10 indicates at least moderate depressive symptoms (88% sensitivity and 88% specificity),^21^ which warrants clinical judgment to determine the necessity of treatment.

Since fatigue is so common after severe COVID-19, assessing depression can be challenging in this population since two of the nine PHQ-9 questions may be influenced by fatigue, and not depression alone, including “*feeling tired or having little energy”* and “*trouble falling or staying asleep, or sleeping too much*.”

To avoid potential overlap in symptoms with fatigue, we also assessed for depressive symptoms using the PHQ-2, which is a briefer two-item screen that assesses the frequency of depressed mood and anhedonia, because it does not include the two PHQ-9 questions described above that may be directly influenced by fatigue, yet shown to retain suitable level of accuracy in a meta-analysis (area under the curve of 0.88 with a sensitivity of 91% and specificity of 67% for cutoff of ≥2).^29^

### Statistical Methods

#### Descriptive Analyses

We used R statistical software version 4.4.1^30^ for all analyses. We described characteristics of the cohort, the FACIT-Fatigue score, and the PHQ-9 using standard descriptive statistics. We first confirmed that the PHQ-9 had excellent reliability in our cohort (a = .90). Next, we explored the relationship between fatigue and depression by describing the distribution of responses to PHQ-9 questions for participants who were not fatigued and fatigued per the FACIT-Fatigue scale. We then conducted bivariate correlation analyses between PHQ-9 and PHQ-2 with FACIT-Fatigue, respectively.

#### MIMIC Model

To assess the influence of fatigue on depression among participants recovering from prolonged severe COVID-19, we used a structural equation modeling approach (SEM) known as the multiple indicators and multiple causes models (MIMIC). This method allows us to better understand complex relationships between overlapping symptoms, such as fatigue and depression, which can be difficult to differentiate with traditional regression methods. A MIMIC model allows us to assess if and how fatigue affects depression and whether the overlap of symptoms might lead to misinterpretation of the severity of depression in survivors of severe COVID-19, by handling multiple related symptoms simultaneously while accounting for errors in how we measure these symptoms.^31^ In other words, the MIMIC model can estimate both observed symptoms (PHQ-9 items) and unobservable factors that underly depression severity. In our study, the MIMIC model identifies the extent to which fatigue directly affects specific depression symptoms (e.g., feeling tired) plus indirect effects through overall depression (as a latent factor that is inferred but not directly observed from PHQ-9 items).^32^ Our MIMIC model schematic is depicted in **Figure 1**.

**Figure 1.**
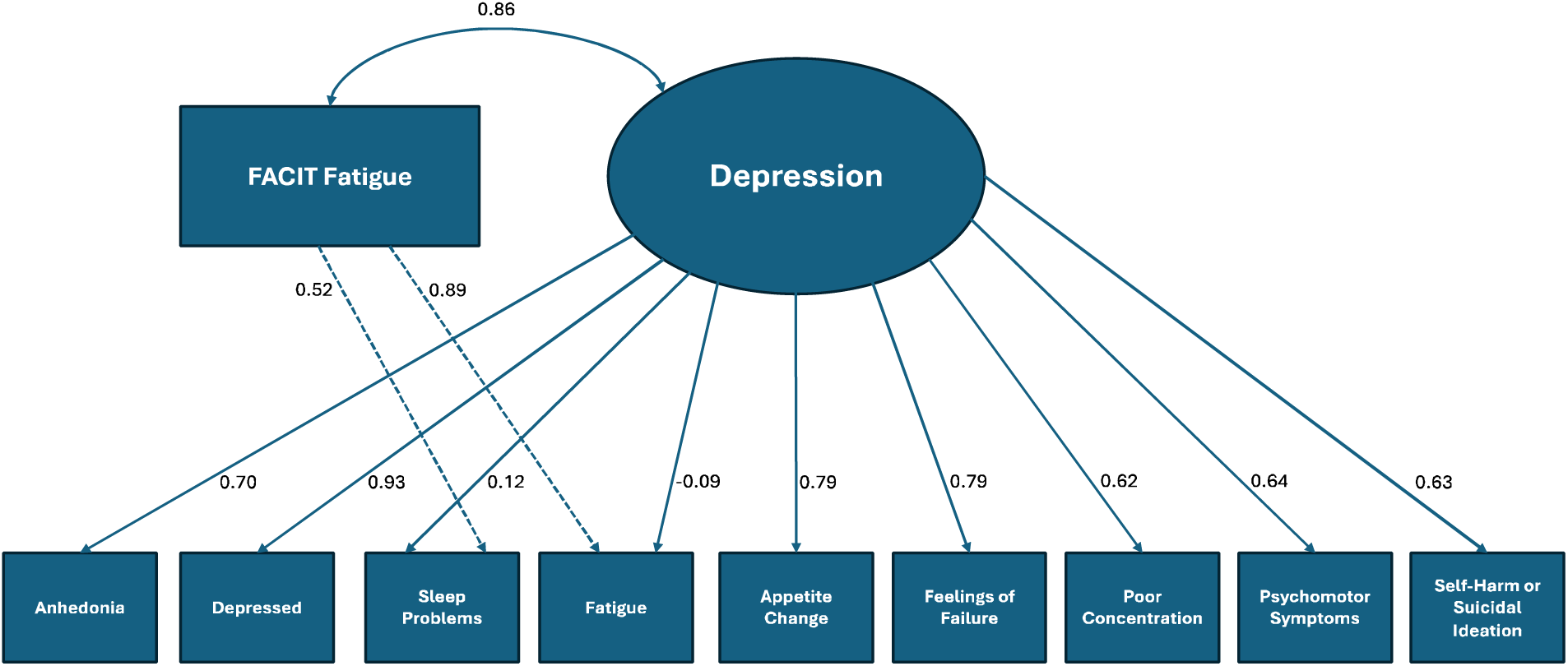
Model Schematic Between Fatigue and PHQ-9.

From our MIMIC model, we estimated standardized factor loadings to assess how strongly each PHQ-9 questionnaire item is linked to the latent depression factor, where higher loadings mean that the item is a more robust indicator (e.g., if the ‘feeling down’ item has a high loading, it is a strong sign of depression). Standardized factor loadings enabled us to compare the strength of each item’s association with the underlying depression construct on a common scale. We then conducted Differential Item Functioning (DIF) to estimate the direct effects of fatigue, which are the immediate impact of the FACIT-fatigue scale on the two a priori selected PHQ-9 questions described above, and the indirect effects of FACIT-fatigue scale, which is the effect of fatigue on the latent construct of depression, which, in turn, affects other depression symptoms. DIF analysis allows us to determine whether PHQ-9 items function differently for individuals with varying levels of fatigue; that is, the item may not measure depression in the same way for people with high fatigue compared to those with low fatigue. By adjusting for DIF pathways, we can more accurately estimate the impact of depression by minimizing the confounding influence of fatigue.^33^

Several criteria were used to assess the fit of our MIMIC model.^34^ We used the Root Mean Square Error of approximation (RMSEA), where values below .05 indicate a good fit, and values up to .08 represent a reasonable error of approximation. Additionally, we tested our model using the Standardized Root Mean Squared Residual (SRMR), which uses a conventional threshold of .08, to determine if the model’s standardized residuals are within an acceptable range. Lastly, we assessed the fit of the model using the comparative fit index (CFI), and Tucker–Lewis index (TLI), which are incremental fit indices using a conventional threshold of .95.

#### PHQ-7 Depression Score

In addition to the PHQ-2 score described above, we conducted an exploratory analysis of a derived simplified PHQ-7 score that excluded the two depressive symptoms of ‘tired’ and ‘sleep’ that were influenced by fatigue in our MIMIC model. We rescaled the score and cutoff to match a PHQ-9 in two different ways. First, we rescaled the PHQ-7 score and cutoff to match the PHQ-9 score and cutoff (e.g., PHQ-7 score of 8 or more equated to at least moderate depression). Second, we matched the distribution of the PHQ-7 to that of the PHQ-9 using a density function and because findings were similar, we herein present the former simpler rescaling approach.

## RESULTS

### Participant Characteristics

Among the 156 eligible participants from the parent RAFT COVID cohort, 82 (52.6%) completed the depression and fatigue surveys at one-year (because FACIT-fatigue was added midway during the study). The median age was 65 years (IQR 60-74), 30.5% were female, 26.8% were non-White, and 59.3% had less than a college education (**Table 1**). Most participants were married (74.4%), employed (70.0%), and self-reported being in good health prior to their COVID-19 illness (63.0%). A minority reported a history of depression (12.2%), anxiety (7.3%), or PTSD (2.4%). Participants had prolonged, severe COVID-19 illness with a median total hospital length of stay of 56 days (IQR 42-75), with two-thirds (67.1%) receiving prolonged mechanical ventilation.

**Table 1.** Participant Characteristics by Availability of 1-Year Fatigue & Depression Assessments.

| Characteristics | Available<br>(N=82) | Unavailable<br>(N=74) |
| --- | --- | --- |
| **Sociodemographics** |  |  |
| Age, median years (IQR) | 65 (60-74) | 64 (55-70) |
| Women | 25 (30.5%) | 35 (47.3%) |
| Race/ethnicity |  |  |
| White, non-Hispanic | 60 (73.2%) | 48 (64.9%) |
| Black | 13 (15.9%) | 22 (29.7%) |
| Latino | 5 (6.1%) | 3 (4.1%) |
| Other | 4 (4.9%) | 1 (1.4%) |
| Education less than college | 48 (59.3%) [n=81] | 41 (55.4%) |
| Social ladder ≤ 5 of 10, % | 23 (28.8%) [n=80] | 24 (33.8%) [n=71] |
| Married | 61 (74.4%) | 46 (62.2%) |
| Employed part- or full-time | 50 (70.0%) | 52 (71.2%) [n=73] |
| **Self-Reported Health Before Illness** |  |  |
| In good prior health <sup>a</sup> | 51 (63.0%) [n=81] | 44 (59.5%) |
| Depression history | 10 (12.2%) | 12 (16.2%) |
| Anxiety history | 6 (7.3%) | 10 (13.5%) |
| PTSD history | 2 (2.4%) | 5 (6.8%) |
| **Severity of the COVID-19 illness** |  |  |
| Total hospital length of stay, median days (IQR) | 56 (42-75) [n=75] | 59 (45-83) [n=72] |
| Mechanical ventilation | 51 (67.1%) [n=76] | 64 (87.7%) [n=73] |
| Mechanical ventilation days, median (IQR) | 34 (14-44) [n=49] | 23 (15-36) [n=57] |
| Tracheostomy | 32 (42.7%) [n=75] | 31 (42.5%) [n=73] |
| **Self-Reported Health at 1 Year Follow-up** |  |  |
| Fatigued (FACIT-Fatigue ≤ 43) | 50 (61.0%) | - |
| FACIT-Fatigue, median (IQR) | 39.5 (31-47) | - |
| Moderate depression (PHQ-9 ≥ 10) | 13 (15.9%) | 17 (23.0%) |
| PHQ-9, median (IQR) | 3 (0-8) | 6 (1-9) |
| Moderate anxiety (GAD-7 ≥ 10) | 15 (18.3%) | 14 (19.2%) [n=73] |
| PTSD (IES-R ≥ 1.6) | 11 (13.4%) | 16 (22.2%) [n=72] |
| Cognitive impairment (TICS ≤ 30) | 15 (18.3%) | 12 (16.2%) |
| ≥1 ADL difficulty | 31 (37.8%) | 36 (48.6%) |
| ≥1 ADL dependence | 10 (13.4%) | 12 (16.2%) |
| Mobility difficulty | 45 (54.9%) | 43 (58.1%) |
| Mobility impairment | 10 (12.2%) | 15 (20.3%) |
| Shortness of breath (mMRC ≥ grade 2) <sup>b</sup> | 37 (45.1%) | 38 (51.4%) |
| ≥1 persistent PICS impairment | 56 (68.3%) | 44 (59.5%) |
| EQ-5D quality of life, median (IQR) | 0.67 (0.58-0.76) | 0.66 (0.54-0.74) |
| Returned to usual health | 11 (13.4%) | 22 (30.1%) [n=73] |
| Returned to work if previously employed | 32 (64.0%) | 29 (55.8%) |

*Abbreviations:* PTSD; post-traumatic stress disorder; FACIT-Fatigue, Functional Assessment of Chronic Illness Therapy-Fatigue; PHQ-9, Patient Health Questionnaire-9; GAD7, Generalized Anxiety Disorder-7; IES-R, Impact Event Scale-Revised; TICS, Telephone Interview of Cognitive Status; ADL, activity of daily living; mMRC, modified Medical Research Council; PICS, post-intensive care syndrome
<sup>a</sup> Good prior health was defined as having no baseline ADL or mobility impairment, breathlessness (mMRC $>1$ ), dementia, psychiatric disorder (depression, anxiety, PTSD), or other serious comorbidity
<sup>b</sup> mMRC grade of 2 (0-4 range) is defined as walking slower than people of the same age because of dyspnea or stop for breath when walking at their own pace.

One year after hospitalization, 61.0% were classified as having fatigue per the FACIT-Fatigue scale and 15.9% as having at least moderate depression per the PHQ-9. About two-thirds reported a persistent PICS impairment (68.3%) and returned to work (64.0%), but only a minority returned to usual pre-illness health (13.4%). More details on self-reported health at one year are shown in **Table 1**.

Participants of the RAFT COVID study who were not included in this sub-study for missing the fatigue assessment were somewhat similar except more were women (47.3%), black (29.7%), on mechanical ventilation during the hospital (87.7%), and at one-year had greater prevalence of moderate-severity depression (23.0%) and functional impairments yet returned to their usual health more often (30.1%). Missing data for both groups was minimal, as shown in Table 1.

### Relationship between Fatigue and Depression: Descriptive Analyses

Overall, participants experiencing fatigue generally reported more severe symptoms of depression for all nine PHQ-9 items compared to those without fatigue (**Table 2**), highlighting the significant overlap between symptoms of fatigue and depression in this population.

**Table 2.** Responses to PHQ-9 questions Stratified by Fatigue.

| PHQ-9 Items | Responses <sup>a</sup> |  |  |  |  |  |  |  |
| --- | --- | --- | --- | --- | --- | --- | --- | --- |
|  | No Fatigue (n=33) |  |  |  | Fatigue (n=49) |  |  |  |
|  | 0 | 1 | 2 | 3 | 0 | 1 | 2 | 3 |
| Disinterest <sup>b</sup> | 32 | 0 | 0 | 1 | 17 | 19 | 7 | 6 |
| Depressed <sup>b</sup> | 31 | 2 | 0 | 0 | 23 | 15 | 6 | 5 |
| Sleep problems <sup>c</sup> | 28 | 3 | 1 | 1 | 8 | 25 | 3 | 13 |
| Tired <sup>c</sup> | 31 | 2 | 0 | 0 | 3 | 20 | 13 | 13 |
| No Appetite | 32 | 1 | 0 | 0 | 24 | 15 | 4 | 6 |
| Feel Bad (Failure) | 30 | 3 | 0 | 0 | 27 | 13 | 2 | 7 |
| Can't Concentrate | 28 | 4 | 0 | 1 | 21 | 16 | 6 | 6 |
| Move Slow/Restless | 31 | 1 | 0 | 1 | 34 | 6 | 3 | 6 |
| Suicidal | 33 | 0 | 0 | 0 | 41 | 4 | 2 | 2 |

Abbreviations: PHQ, Patient Health Questionnaire
<sup>a</sup> Responses for each PHQ-9 item are self-rated by participants on a 4-point Likert scale coded as 0 for “not at all”, 1 for “several days”, 2 for “more than half the time”, and 3 for “nearly every day” during the past two weeks.
<sup>b</sup> these two items comprise the PHQ-2
<sup>c</sup> these items are theorized to be influenced by fatigue in our conceptual model

Fatigue was significantly correlated with the PHQ-9 (*r=*.87, *p*<.001) and, to a lesser extent, with the PHQ-2 (*r=*.76, *p*<.001), which did not include the two questions pertaining to fatigue.

### Influence of Fatigue on Depression: Findings from the MIMIC Model

#### Parameter Estimates and Decomposition

Our base PHQ-9 model without any DIF pathways confirmed that the nine PHQ-9 items strongly loaded on the underlying depression construct in our cohort (**Table 3**). From the MIMIC model which included the two DIF pathways, fatigue had significant standardized direct effects on difficulty sleeping (λ = .52, *p*<.001) and tiredness (λ = .89, *p*<.001), with a decrease in the standardized effects of these two PHQ-9 items on the depression construct (**see Figure 1 and Table 3**).

**Table 3.**
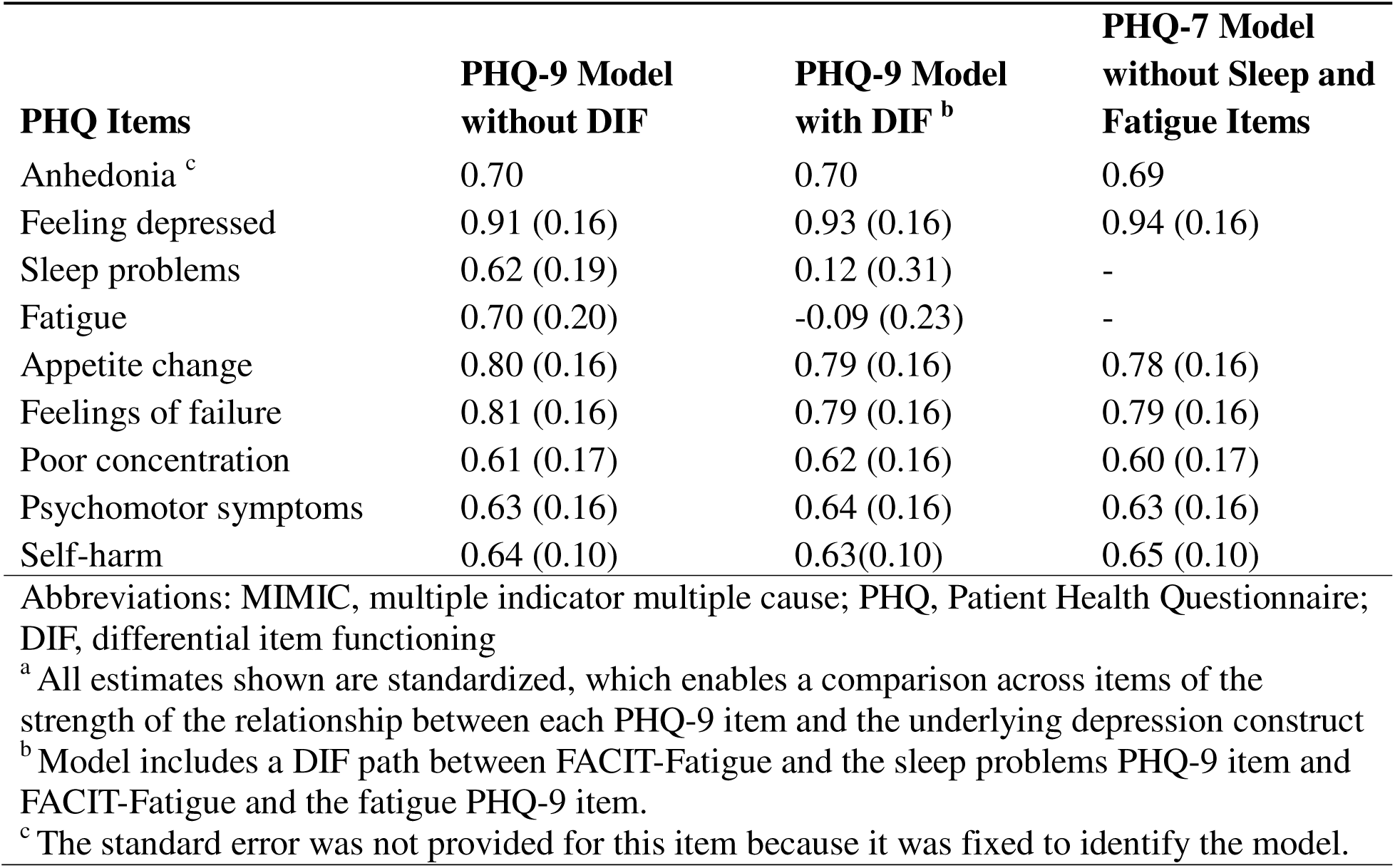
MIMIC Model Estimates for Depression ^a^.

Indirect effects were estimated to understand how fatigue influenced other depressive symptoms via the latent depression construct. The indirect effects of fatigue on sleep (λ = .10, *p* = .53) and on tiredness (λ = -0.08, *p*=.54) were low through the latent depression factor.

The total standardized effect from the FACIT-Fatigue scale on depression in our model, which combines both direct effects on fatigue-related symptoms (e.g., tiredness and sleep difficulties) and indirect effects on the broader depression construct, suggests fatigue contributes to depression severity, but was not statistically significant (λ = 1.44, *p=*.88).

#### Model Fit and Estimation

The fit indices of the MIMIC model were mixed. The RMSEA was .153 (90% CI .114, .192), slightly above .08 value which represents a reasonable error of approximation.^34^ In contrast, the SRMR was .070, suggesting that the model’s standardized residuals were within an acceptable range. Additionally, the CFI (.874) and TLI (.832) were below the conventional threshold of .95, although close to levels commonly seen with small sample sizes, suggesting the model fit is interpretable.^35^

#### PHQ-7 Depression Score

As a companion analysis to the MIMIC model, we examined the impact of using a simplified PHQ-7 score that omitted the two PHQ-9 items (tiredness and sleep problems) influenced by fatigue. The PHQ-7 items strongly loaded on the depression construct (**Table 3**). PHQ-7 was strongly correlated with the PHQ-9 (*r=*.97, *p*<.001; **see Figure 2**) and with fatigue (*r=*.82, *p*<.001), but to a lesser extent than the PHQ-9.

**Figure 2.**
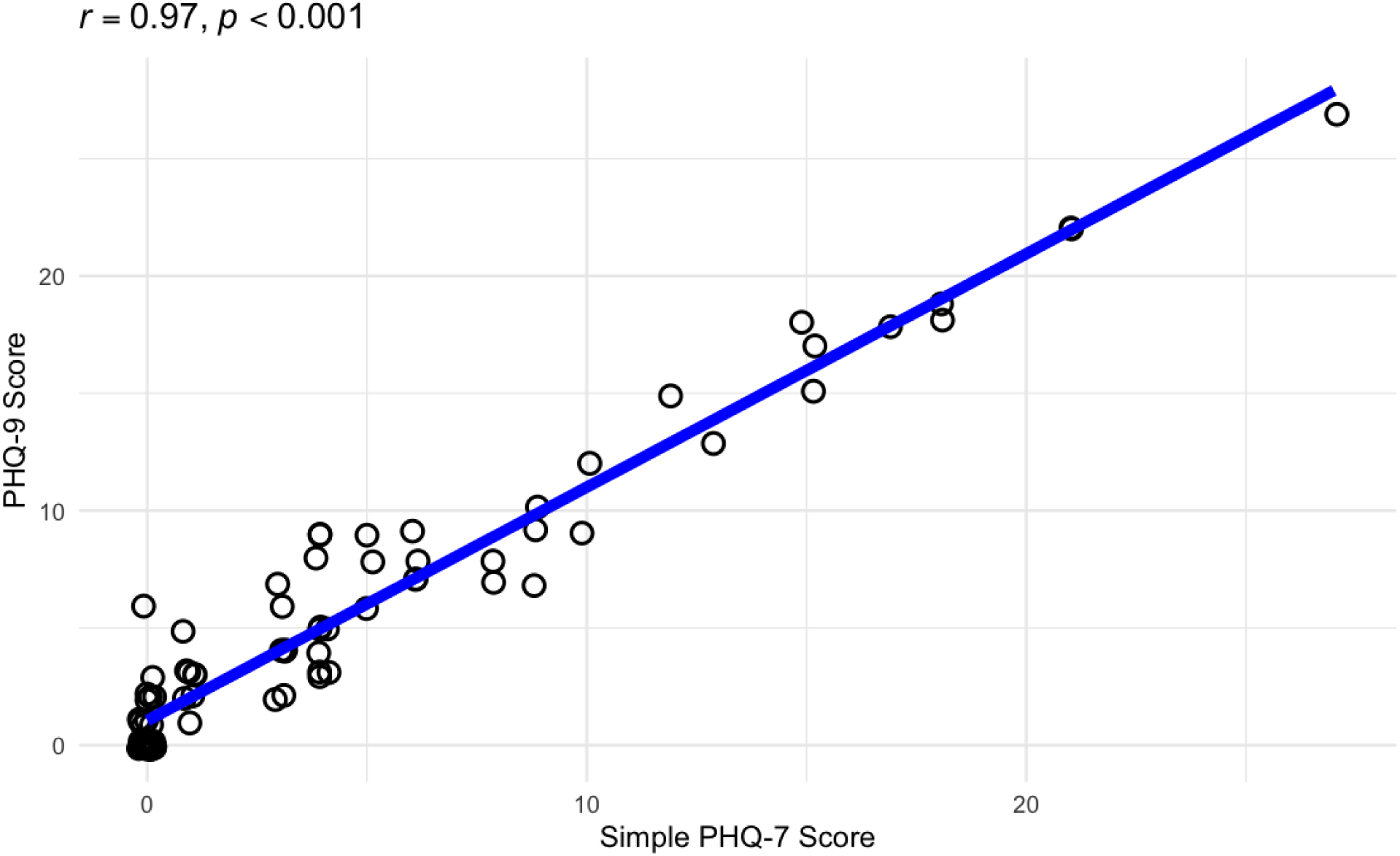
PHQ-9 vs re-scaled PHQ-7 scores.

The rescaled PHQ-7 score was modestly lower than the PHQ-9 in our cohort, particularly in the lower range of the scale (e.g. less depressive symptoms) where PHQ-9 tended to overestimate scores relative to PHQ-7. Among those who were not fatigued, no participants were considered depressed by either the PHQ-9 (median score of 0, IQR 0-2) or the PHQ-7 (median score of 0, IQR 0-1). Among those who were fatigued, the rescaled PHQ-7 (median score of 4.5 IQR 1.50-9.75) was lower than the PHQ-9 (median score of 7, IQR 4-9.75), but both classified 15.9% of participants as depressed.

## DISCUSSION

In this national multicenter cohort of survivors of prolonged severe COVID-19 illness, we found a significant overlap between symptoms of fatigue and depression that can complicate the accurate diagnosis and management of depressive symptoms. We present two complementary analyses showing that the PHQ-9 can overestimate depression severity in the presence of fatigue because it includes two items related to sleep disturbances and fatigue that may better capture symptoms of fatigue rather than depression. First, we found higher correlation between PHQ-9 scores and fatigue levels compared to the correlation of fatigue with either the PHQ-2 or the derived PHQ-7, each of which omit these two items. Second, our MIMIC model further clarified these relationships by showing that fatigue directly influenced depression, mostly driven by the direct effects of the FACIT-Fatigue scale through these two PHQ-9 items on sleep and fatigue. These findings underscore the importance of confirming depression in the presence of fatigue among individuals with PICS after severe COVID-19 and careful selection of screening tools for depression in PICS that may not overestimate depression severity that potentially can lead to misdiagnosis or overtreatment.

While there are no other studies to our knowledge that have disentangled fatigue from depression in PICS or after severe COVID illness, our findings align with the broader body of literature for other illnesses where symptoms that overlap with depression.^23–25^ For instance, fatigue from multiple sclerosis strongly overlapped with depression symptoms and that accounting for this overlap led to lower depression scores.^23^ Our study similarly identified significant overlap between fatigue and pertinent depression symptoms among individuals with PICS after severe COVID-19 infection, reinforcing the need for careful selection and adjustment of screening tools in populations where fatigue is prevalent.

The implications of these findings are particularly relevant for clinical practice. In survivors of critical illness, the PHQ-9 may not be the most accurate tool for screening depression given the overlap with fatigue. The inclusion of fatigue-related items in the PHQ-9 can lead to an overestimation of depression severity, which in turn could result in unnecessary or inappropriate treatment interventions. Instead, the use of PHQ-2, or potentially the ad hoc PHQ-7 measure pending validation, may be the preferred screening tool since these scales omit the two items strongly influenced by fatigue. This approach could help in better identifying depression symptoms that may be more amenable to depression treatment with therapy and medications.

Our study has certain limitations. First, our cohort was relatively small. However, we included individuals from a diverse, national, multicenter cohort and nonetheless identified large and significant effects between fatigue and depression. Larger cohorts are needed to both confirm these results and provide more precise estimates of the impact of fatigue on reclassification of depression severity, especially given inconsistent model fit indices, which may reflect limited power rather than being substantively misfit. Second, our findings are specific to PHQ questionnaires. The overlap with fatigue may not also be true of other depression screening tools, such as the Hospital Anxiety and Depression Scale (HADS), which is also commonly used among critical illness survivors.^36^ Although we cannot directly comment on the utility of HADS, there may also be significant overlap between fatigue from PICS and the HADS item, “*I feel as if I slowed down*,” which should be explored in future research. Third, the PHQ-7 findings should be considered exploratory as its diagnostic accuracy for depression requires prospective validation before clinical use. Finally, our study focuses on PICS from severe COVID-19. Whether this is true of critical illness survivors from other illnesses should be explored in future research.

In conclusion, our study contributes to the growing body of literature on the intersection of fatigue and depression, both of which are common to PICS among survivors of severe COVID-19. By highlighting the significant overlap between fatigue and depression, we provide novel evidence to preferentially consider the use of the PHQ-2, which omits the two items on sleep disturbances and fatigue, instead of the PHQ-9 to screen for depression to avoid overestimating depression severity and potentially unnecessary treatment.

## Data Availability

All data produced in the present study are available upon reasonable request to the authors

## ACKNOWLEDGMENTS

We sincerely thank the study participants for sharing their experiences with us. We thank the participating hospitals and personnel who contributed their effort in kind. We thank the National Association of Long Term Hospital Executive Leadership team for their help in coordinating the execution of the study, including Dr. Ed Prettyman (CEO), Mr. Lou Little (past CEO), Dr. John Votto (CMO). NALTH leaders did not receive compensation for their contributions. We thank the following research assistants who conducted data collection: Giselle Aguayo Ramirez, Karen Reyes, and Daisy Brambila. They received financial compensation for their contributions.

## COMPLIANCE WITH ETHICAL STANDARDS

This work was supported by grants from the NIH/NIA (K23AG052603), the UCSF Research Evaluation and Allocation Committee (REAC) (Carson and Hampton Research Funds), and the National Association of Long Term Hospitals (NALTH). We have no conflicts of interest to disclose, financial or otherwise. The University of California, San Francisco Institutional Review Board approved the RAFT COVID study (#20-31060). All study participants provided informed consent. All research procedures were followed in accordance with the ethical standards of the responsible committee on human experimentation and with the Helsinki Declaration of 1975.

